# A Comparative COVID 19 Characterizations and Clinical Course Analysis between ICU and Non ICU Settings

**DOI:** 10.1101/2020.10.07.20208389

**Authors:** Amit Patel, Parloop Bhatt, Surabhi Madan, Nitesh Shah, Vipul Thakkar, Bhagyesh Shah, Rashmi Chovatia, Hardik Shah, Minesh Patel, Pradip Dabhi, Aditi Nanavati, Milan Chag, Keyur Parikh

## Abstract

**Objective:** With COVID-19 pandemic severely affecting India and Ahmedabad city being one accounting for half COVID cases, objective was to determine disease course and severity of in patients at a COVID care hospital.

**Design:** A Clinical trial registry of India registered observational study (CTRI/2020/05/025247).

**Setting:** **C**ertified COVID hospital located in Ahmedabad, Gujarat, India.

**Participants:** 549 COVID positive patients hospitalized between 15 ^th^ May to 10 ^th^ August, 2020 and treated in ICU and non ICU settings.

**Main Outcome Measure:** Comparative analysis of demographic, clinical characteristics, investigations, treatment, complications and outcome of COVID patients in ICU and non ICU settings.

**Results:** Of the 549 hospitalized COVID positive patients, 159 were admitted in ICU during disease course while 390 had ward admissions. Overall median age was 52 (1-86) years. The ICU group was older (>65years), with associated comorbidities like hypertension and diabetes (p<0.001); higher proportion of males (79.25%); with dyspnea as a major clinical characteristic and consolidation in lungs as a major radiological finding as compared to ward patients. C - reactive protein, D-Dimer and Ferritin were higher in ICU patients. Overall 50% females depicted elevated Ferritin levels. Steriods(92.45%)and tocilizumab (69.18%) were more frequently used for ICU patients. Remdesivir was prescribed to both ICU and non ICU patients. Favirapir was also a line of treatment for 25% of ICU patients. Convalescent plasma therapy was given to 7 ICU patients. Complications like acute kidney injury (13.84%), shock (10.69 %), sepsis and encephalopathy were observed in ICU patients. Overall mortality rate was 5.47 % with higher mortality among males in comparison to females (p<0.0001).

**Conclusion:** About 29% of overall patients required ICU admission that was commonly elderly males. Chances of ICU admission were higher with baselines comorbidities (1.5 times) and dyspnea (3.4 times) respectively. A multi-specialty COVID care team and updated treatment protocols improves outcomes.

## Introduction

The global COVID-19 pandemic caused by severe acute respiratory syndrome coronavirus 2 (SARS-CoV-2) has severely affected India. As reported on 6^th^ October 20202, with 6,685082 corona virus cases and 103600 reported deaths, India has the largest number of confirmed cases in Asia ^[1]^ and has the second highest number of confirmed cases in the world after the United States. ^[1]^ Gujarat is one of Indian states severely affected by the corona virus, with 123638 cases reported so far ^[2]^ of which Ahmedabad the economic city of Gujarat reports more than 37,000-covid cases.^[3]^ The government of Gujarat partially locked the city of Ahmedabad on 21^st^ March ^[4]^ and implemented various policies to curb the pandemic.^[5]^ Despite the pressing need for evidence to inform such key decisions, data remain limited on COVID-19 in India. Patient characteristics, illness course, practice patterns, resource utilization, morbidity, and mortality associated with COVID-19 have been characterized in only limited samples which is scattered. Characteristics of Gujarat state patients are beginning to be enumerated with limited data on hospitalized patients, including the critically ill; therefore, we sought to characterize the disease course of adult COVID-19 patients admitted at Care Institute of Medical Sciences (CIMS hospital), Ahmedabad, Gujarat, India. We provide a detailed description of demographic data, comorbidities, presenting symptoms, clinical course, hospital complications, patient outcomes, and mortality of 549 COVID positive patients hospitalized and treated. The analysis also compares the clinical characteristics and its outcomes between patients admitted in ICU and non ICU units during the course of disease.

## Methodology

### Data source and study sample

CIMS hospital is a tertiary care medical center (JCI, NABH accredited) with 350 adult beds serving a diverse, high acuity patient population in Ahmedabad, India. A separated WEST wing of the hospital was designated as a COVID hospital to treat 100 patients. The present data was documented and collected from the medical records department to identify patients with laboratory-confirmed COVID-19 infection, as represented by a positive SARS-CoV-2-RT-PCR test. We performed ongoing retrospective manual data abstraction from the records of 549 patients with COVID-19 who received inpatient care at CIMS hospital from 15^th^ May to 10^th^ August 2020. All hospitalized patient were consented for use of data for research and education. The most common criterion for hospital admission for COVID-19 patients was room air hypoxemia. In the present study as per hospital protocol in line with Indian council of medical research (ICMR) guidelines COVID 19 disease severity has been defined as mild in case of no evidence of breathlessness or hypoxia; moderate with presence of clinical features of dyspnea and or hypoxia, cough, fever including SpO2 between 90 to 94% on room air, respiratory rate more than or equals to 24 per minutes; Severe with clinical signs of Pneumonia including respiratory rate >30 breaths/min, severe respiratory distress, SpO2 <90% on room air.^[6]^ For this manuscript, an ICU bed is defined as one with the capability of providing mechanical ventilation and continuous vital sign monitoring, with staffing by critical care nurses and oversight by intensivists.

### COVID treating and chart review team

Supervised by multiple clinicians including intensivists, clinicians, pulmonologists, anesthetists, infectious disease expert, pathologist, microbiologist, pharmacologist, cardiologist and informaticians, an abstraction team of 9 trained data analytics manually abstracted medical records data in chronological order by admission date. Information from the charts was inputted directly into hospital information management software besides paper medical record forms. Records with missing data or with inconsistent times were reviewed by a second, dedicated quality controller. A random subsample of abstracted data was checked by a second abstractor, typically a clinician, for calibration and consistency. Any conflicting data were resolved by consensus.

Data collected include demographics, comorbidities, presenting symptoms, laboratory and radiographic findings, hospital course, mechanical ventilation, complications (defined as those documented by clinicians in the medical records) such as acute respiratory distress syndrome (ARDS) or acute kidney injury (AKI), and disposition including discharge, death or leave against medical advice (LAMA).

Time of first symptom was recorded based on the patient’s history; if the patient did not or could not give a specific date of first symptom, it was recorded that the patient could only give an approximate time. Data present in the medical record were only included for analysis; no imputation was performed. Laboratory test data were extracted from the in-house laboratory data software validated with daily assessments.

### Data characterization and analysis

Individual records were labeled with the highest level of care a patient received during hospital stay as (1) Ward hospitalized (non-ICU), and (2) ICU admission. Characteristics were stratified by the highest level of care received to date, and 95% confidence intervals (CIs) were recorded for each value. All analyses were performed using STATA, version 11.0. Continuous variables were reported as medians and interquartile ranges (IQR). Means were determined for frequency variables. For statistical significance, measures of effect; odds ratios with 95% CI’s, and a 2-sided p value less than 0.05 were applied. Logistic regression analysis was applied wherever appropriate.

## Results

A total of 1291 patients were tested positive for SARS-CoV-2, and were hospitalized for treatment in intensive care unit (ICU) or non ICU units between 15^th^ May to 10^th^ August 2020. Medical records from each month were randomly picked for analysis. During the first month of COVID treatment i.e May 2020, since the processes were getting established, 25% (55/221) of medical records were collected; June ‘20, 69% (285/415),July ‘20, 35 % (168/474) of data were analyzed. Data of first 10 days of August ‘20 were analyzed limiting the medical records to 22% (41/181). Our cohort includes randomly picked 549 of these 1291 patients. Of these 549 patients, 159(28.96%) patients were treated in ICU while 390 hospitalized patients did not require ICU-level care.

### Baseline characteristics

Table 1 presents a detailed breakdown of baseline characteristics including age groups and gender. Overall median age was 52 (1-86) years of which 72.5 % were males. A one year old child was also tested COVID positive and admitted in non ICU settings. Age greater than 65 years was a contributing parameter for ICU admission (29.56 vs. 21.12 %; p=0.002).Younger patients (18-34 years) had less disease severity (ICU: 5.0 % vs. ward: 22.3%; p<0.0001). There was a male predominance in the overall sample which was more pronounced among ICU patients (79.25% males).

**Table 1.**
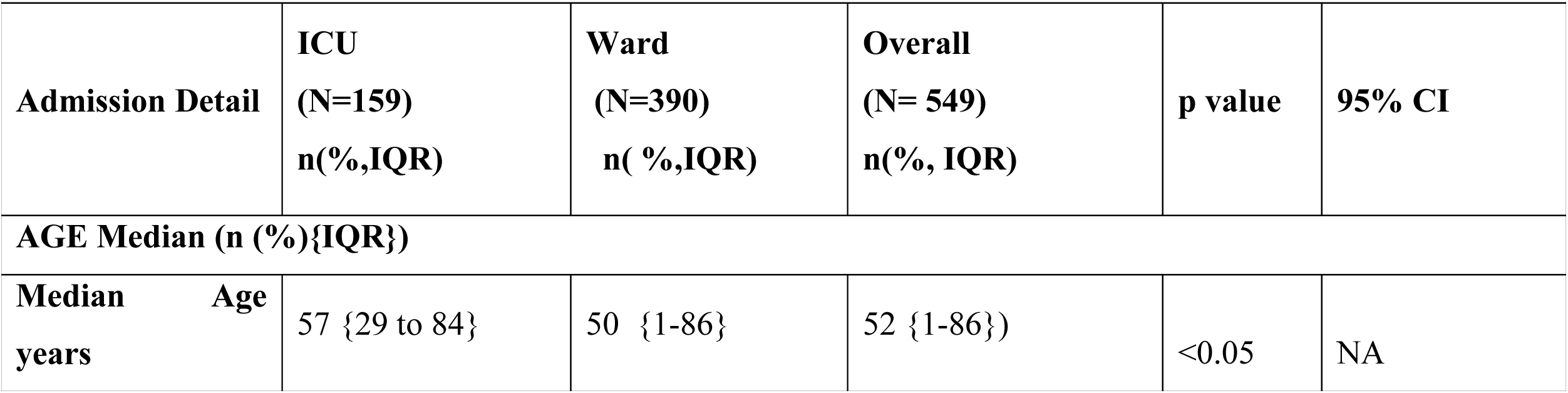
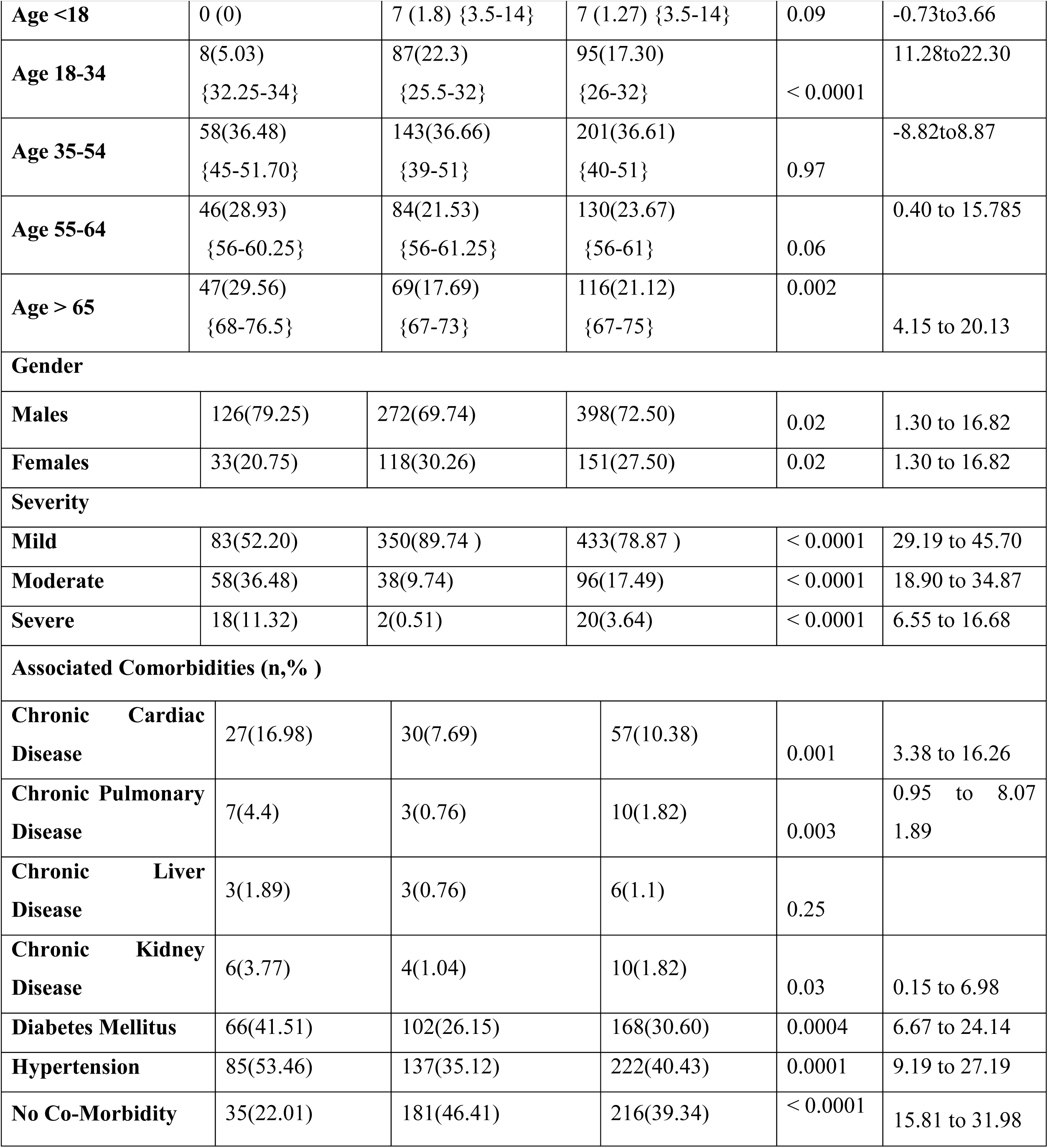
Baseline Characteristic and Associated Comorbidities

As per ICMR guidelines, disease severity was classified based on SpO_2_ and respiration rate. Of the 549 patients, 433 (78.87%) patients had mild disease, 96 (17.49%) had moderate while 20 (3.64%) had severe disease on admission.

### Associated Co morbidities

Hypertension was the most common comorbidity, present in 40.43% of patients, followed by diabetes in 30.6%; cardiac disease (10.38%) (Table 1).Prevalence of comorbidities was significantly higher (p<0.001) in ICU patients as compared to ward patients; hypertension (53.46 vs. 35.12%), diabetes (41.51 vs. 26.15%),cardiac disease (16.98 vs. 7.69%).Other co-morbidities included chronic pulmonary disease(Overall:1.82%; ICU:4.4 % vs. ward 0.76 % ; p = 0.0038), chronic liver disease (Overall:1.10%;ICU:1.89 vs. ward 0.76;p=0.247),chronic kidney disease (Overall:1.82%;ICU:3.77% vs. ward 1.04%;p=0.0306). 39.34 % of patients reported no major comorbidities more so in non ICU settings (p<0.0001).

### Clinical Characteristics

Patients’ most common presenting symptoms were fever (89.61%), cough (58.28%), and shortness of breath (10.74%) (Table 2).Body ache (26.41 %) more so headache was reported in 14.93% of patients. Fatigue/weakness was observed in 27.86 % of patients with 13.11 % reporting sore throat. Dyspnea as a presenting symptom was significantly more common in those who progressed to the ICU (p<0.0001), while non-ICU patients had higher rates of headache (17.44% vs.8.81 %;p=0.01),dysgeusia (7.65 % vs.1.89 %;p=0.009) and anosmia (5.12% vs. 1.26 %;p=0.03).

**Table 2.**
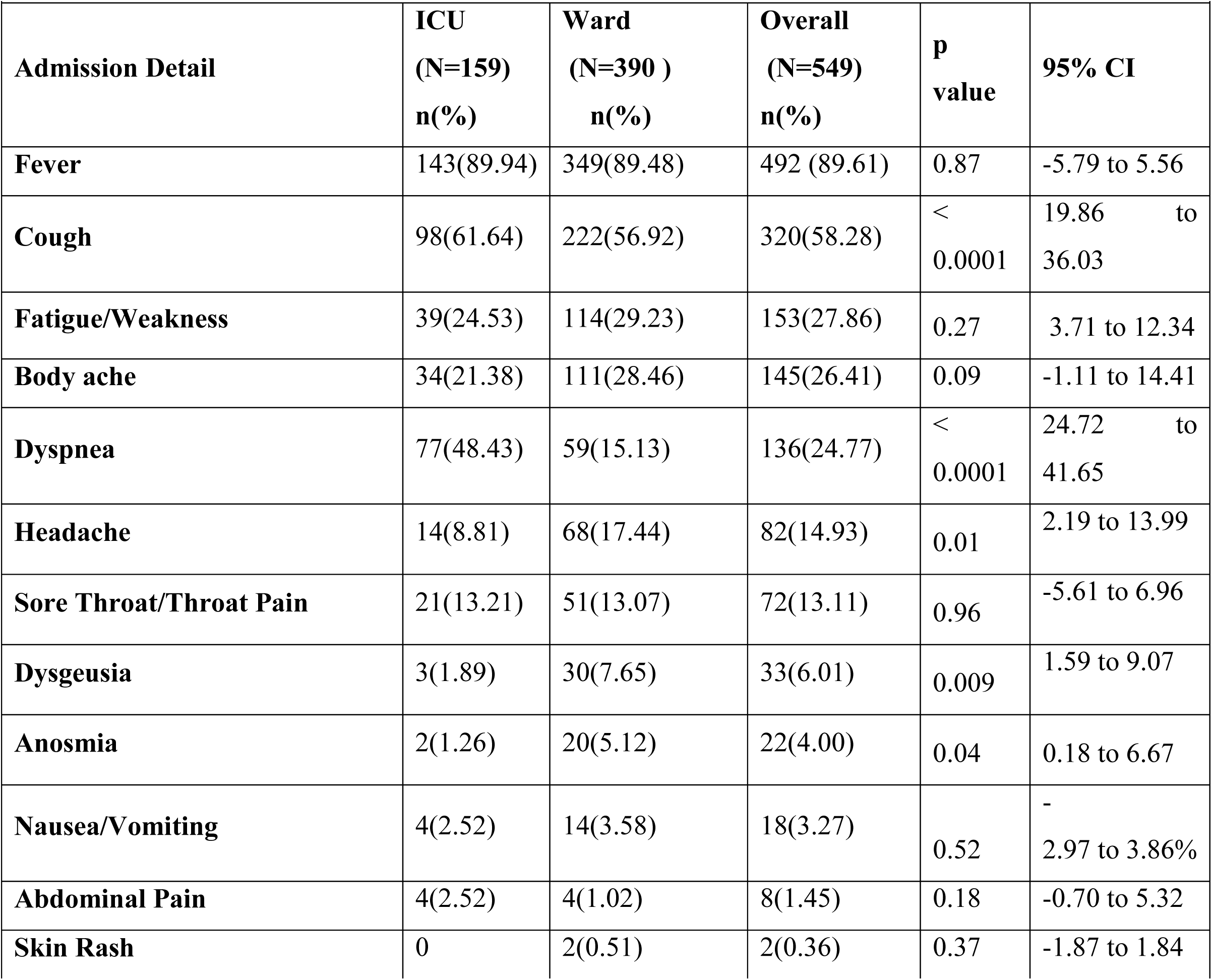
Clinical characteristics

### Laboratory Investigations and Radiological Findings

Although exhaustive laboratory investigations were performed, this manuscript presents inflammatory marker C-reactive protein (CRP),D-DIMER and Ferritin.(Table 3). Nearly 90 % all hospitalized patients had elevated inflammatory marker CRP.D-dimer levels were significantly higher in ICU patients as compared to ward patients. Overall 50 % of admitted females had high levels of ferritin.

**Table 3:** Radiological Findings and Laboratory Investigations

| Table 3 : Radiological Findings and Laboratory Investigations |  |  |  |  |  |  |  |  |  |  |  |
| --- | --- | --- | --- | --- | --- | --- | --- | --- | --- | --- | --- |
| Variable | Ranges | ICU |  | Ward |  | Overall |  | p Value |  | 95% CI |  |
|  |  | (N= 159) |  | (N=390) |  | (N= 549) |  |  |  |  |  |
| Markers (Unit) |  | % |  | % |  | % |  |  |  |  |  |
| CRP (mg/dL) | < 0 | 0 |  | 0 |  | 0 |  | NA |  | NA |  |
|  | Normal :0 to 5 | 8.28 |  | 11.92 |  | 10.9 |  | 0.21 |  | -2.40 to 8.56 |  |
|  | > 5 | 91.72 |  | 88.08 |  | 89.11 |  | 0.21 |  | -2.40 to 8.56 |  |
| D-DIMER (ng/ml) | < 0 | 0 |  | 0 |  | 0 |  | NA |  | NA |  |
|  | Normal :0 to 500 | 42.55 |  | 70.63 |  | 62.47 |  | < 0.0001 |  | 18.99 to 36.65 |  |
|  | > 500 | 57.45 |  | 29.36 |  | 37.53 |  | < 0.0001 |  | 19.00 to 36.66 |  |
| FERRITIN (ng/ml) | Gender | Male | Femal e | Male | Femal e | Male | Femal e | Male | Femal e | Male | Female |
|  | Low | 16.67 | 20 | 8.13 | 6.34 | 10.94 | 9.64 | 0.01 | 0.01 | 1.82 to 16.48 | 1.90 to 30.37 |
|  | Normal: Males :30-400 Females: 15-150 | 51.19 | 30 | 65.11 | 42.85 | 60.55 | 39.76 | 0.008 | 0.18 | 3.55 to 24.11 | 6.21 to 28.55 |
|  | High | 32.14 | 50 | 26.74 | 50.79 | 28.51 | 50.6 | 0.27 | 0.93 | -3.95 to 15.28 | -17.64 to 19.20 |
| Abnormal chest X-ray findings |  | 131(82.38) |  | 178(45.64) |  | 309(56.28) |  | < 0.0001 |  | 28.43 to 43.82 |  |
| Abnormal CT-scan findings |  | 74(46.54) |  | 114(29.23) |  | 188(34.24) |  | 0.0001 |  | 8.39 to 26.16 |  |

Overall 56.28% of patients had abnormal chest X ray findings.82.38% of ICU patients reported consolidation in the lungs. In ward patients although disease severity was less 45.64% reported abnormal chest X ray. With protocol amendments, CT scan was incorporated as a diagnostic tool, wherein ICU patients reported abnormal findings (p=0.001) (Table 3).

### Treatment

Overall, 66.3% of patients received over 48 hours of antibiotic therapy during their stay (most commonly azithromycin) and 63.2% received hydroxychloroquine (Table 4). Both treatments were prevalent in ICU and non ICU patients. Other antibiotics (83.65%) were also offered in ICU as compared to non ICU patients. Steriods(92.45%) and tocilizumab (69.18%) were more frequently used for ICU patients as compared to non ICU patients. Initially methylprednisolone was prescribed but revised protocols allowed use of dexamethasone also. About 27.14% were given antivirals. With availability of remdesivir, it was prescribed to both ICU and non ICU patients. Favirapir was also a line of treatment for 25% of ICU patients. With updated protocols, 7 ICU patients were treated with convalescent plasma as per ICMR guidelines.

**Table 4.** Inpatient Medications

| **Admission Detail** | **ICU (N= 159)**<br>**n(%)** | **Ward (N=390)**<br>**n(%)** | **Overall (N= 549) n(%)** | **p value** | **95% CI** |
| --- | --- | --- | --- | --- | --- |
| **Hydroxychloroquine** | **87(54.72)** | **260(66.66)** | **347(63.20)** | **0.009** | **2.99 to 20.90** |
| **Azithromycin** | **107(67.3)** | **257(65.89)** | **364(66.30)** | **0.75** | **-7.45 to 9.76** |
| **Steroids** | **147(92.45)** | **146(37.43)** | **293(53.36)** | **< 0.0001** | **47.89 to 60.66** |
| Methylprednisolone | 143 (89.94) | 134 (91.78) | 277 (50.45) | 0.49 | 3.07 to 7.96 |
| Dexamethasone | 3(1.89) | 11(7.53) | 14(2.55) | 0.011 | 1.48 to 8.93 |
| Hydrocortisone | 1(0.63) | 0 | 1(0.18) | 0.11 | -0.47 to 3.47 |
| **Anti-Virals** | **56(35.22)** | **93(23.84)** | **149(27.14)** | **0.006** | **3.07 to 20.03** |
| Remdesivir | 41(73.21) | 85(91.40) | 126(84.56) | < 0.0001 | 11.14 to 25.94 |
| Favipiravir | 14(25) | 6(6.45) | 20(13.42) | < 0.0001 | 11.81 to 26.09 |
| Acyclovir | 1(1.79) | 2(2.15) | 3(2.01) | 0.79 | -3.26 to 2.66 |
| **Antibiotics** | **133(83.65)** | **116(29.74)** | **249(45.35)** | **< 0.0001** | **45.85 to 60.47** |
| Amoxicillin | **9 (3.46)** | **4 (2.48)** | **13 (3.08)** | **0.53** | **-1.83 to 5.24** |
| Ampicillin | **1 (0.38)** | **0** | **1 (0.23)** | **0.22** | **-0.65 to 3.05** |
| Cefixime | **144 (55.38)** | **132 (81.98)** | **276 (65.55)** | **< 0.0001** | **18.03 to 35.11** |
| Polymixin-B | **13 (5.0)** | **0** | **13 (3.08)** | **< 0.0001** | **2.36 to 9.57** |
| Piperacillin-Tazobactam | **9 (3.46)** | **2 (1.24)** | **11 (2.61)** | **0.08** | **-0.32 to 6.39** |
| Meropenem | **46 (17.69)** | **8 (4.96)** | **54 (12.82)** | **< 0.0001** | **6.95 to 19.62** |
| Cotrimoxazole | **3 (1.15)** | **0** | **3 (0.71)** | **0.03** | **-0.14 to 4.30** |
| Other | **35 (13.46)** | **15 (9.31)** | **50 (11.87)** | **0.15** | **-1.38 to 10.81** |
| **Tocilizumab** | **110(69.18)** | **20(5.12)** | **130(23.6)** | **< 0.0001** | **56.04 to 70.94** |
| **Convalescent Plasma** | **7 (4.40)** | **0** | **7 (1.27)** | **< 0.0001** | **1.94 to 8.80** |

### Hospital Complications and Length of Stay

Complication rates were higher in ICU patients with 13.84 % developing acute kidney injury (AKI); 10.69 % developed shock;8.18% sepsis and 5.03 % encephalopathy. Non ICU patients had an uneventful hospital stay (Table 5). Of the 549 patients, 502 (92.28%) recovered with 8.5days of average length of hospital stay. ICU patient’s disease course averaged to 11 days; however one patient was discharged at 69 days. Ward patients had a shorter disease course (Table 5).

**Table 5.**
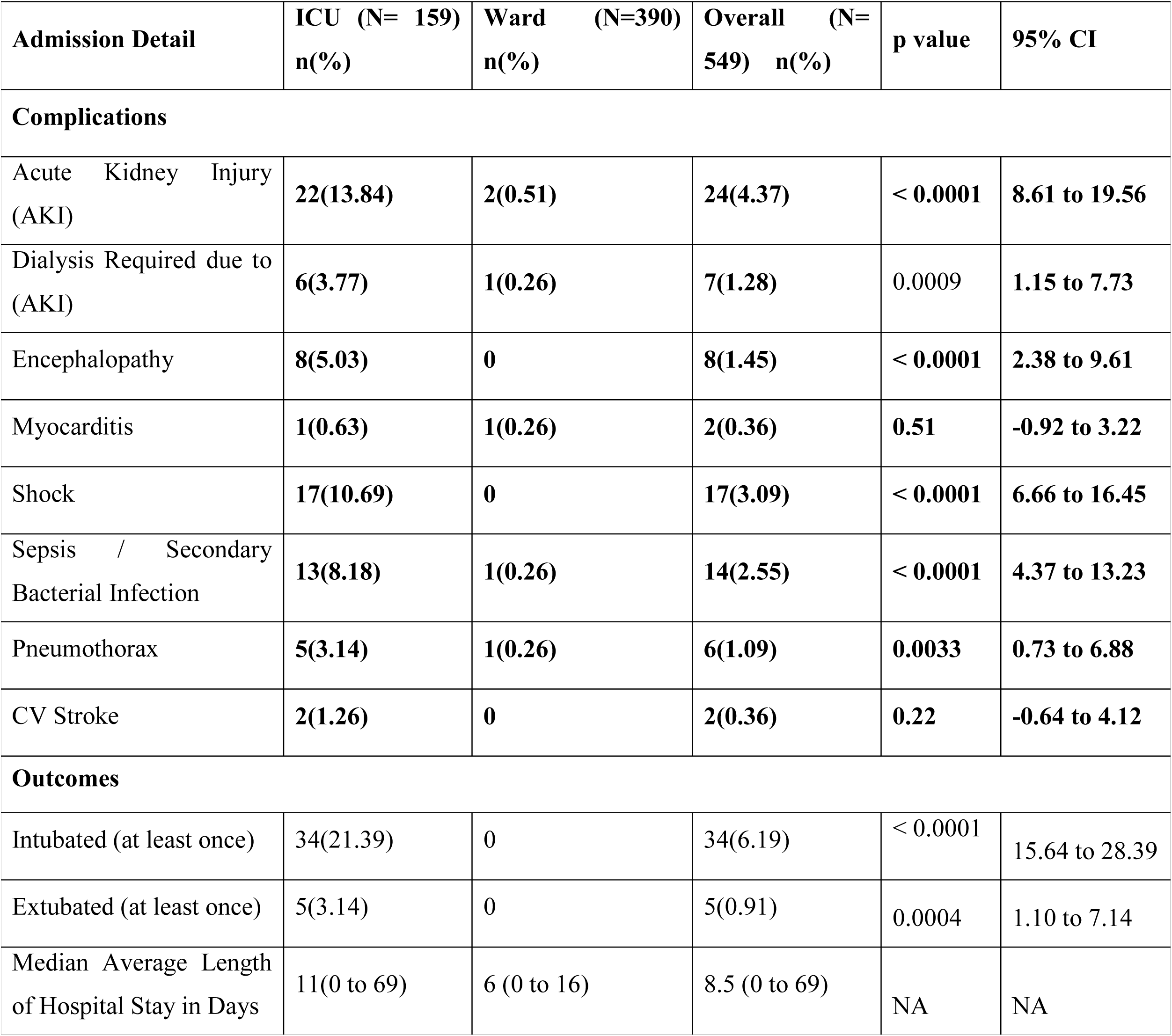
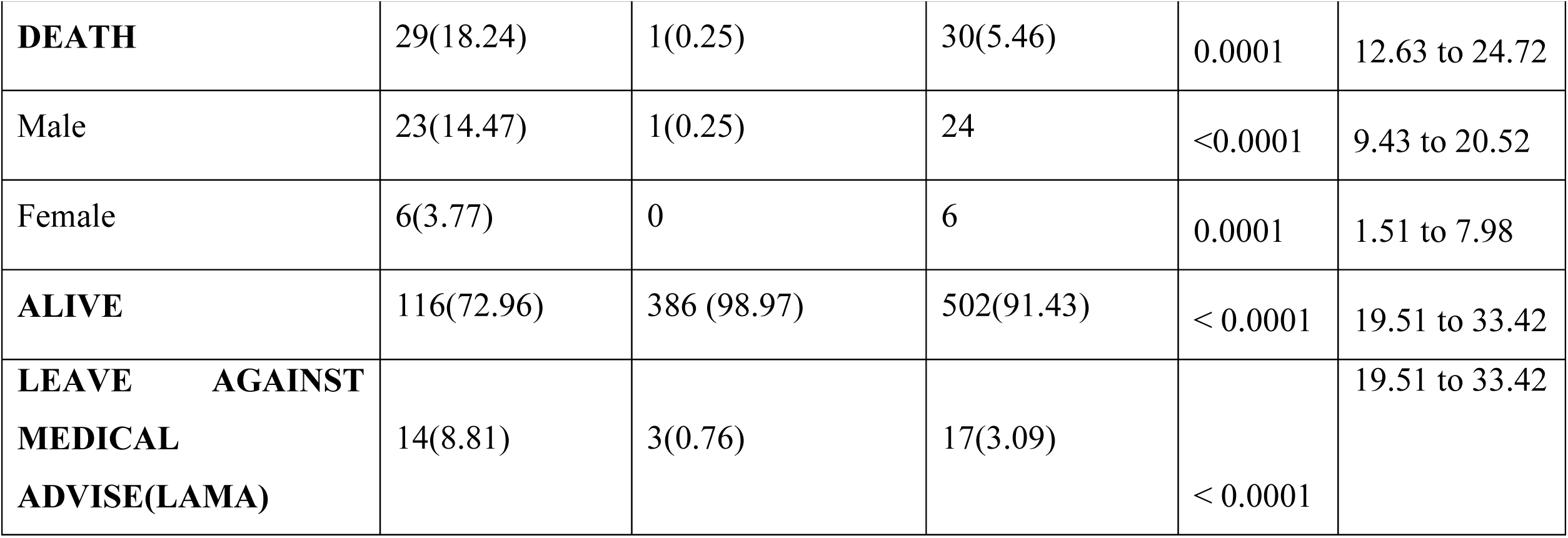
Inpatient Complications and Outcomes

| **Admission Detail** | **ICU (N= 159)**<br>**n(%)** | **Ward (N=390)**<br>**n(%)** | **Overall (N= 549) n(%)** | **p value** | **95% CI** |
| --- | --- | --- | --- | --- | --- |
| **Complications** |  |  |  |  |  |
| Acute Kidney Injury (AKI) | **22(13.84)** | **2(0.51)** | **24(4.37)** | **< 0.0001** | **8.61 to 19.56** |
| Dialysis Required due to (AKI) | **6(3.77)** | **1(0.26)** | **7(1.28)** | 0.0009 | **1.15 to 7.73** |
| Encephalopathy | **8(5.03)** | **0** | **8(1.45)** | **< 0.0001** | **2.38 to 9.61** |
| Myocarditis | **1(0.63)** | **1(0.26)** | **2(0.36)** | **0.51** | **-0.92 to 3.22** |
| Shock | **17(10.69)** | **0** | **17(3.09)** | **< 0.0001** | **6.66 to 16.45** |
| Sepsis / Secondary Bacterial Infection | **13(8.18)** | **1(0.26)** | **14(2.55)** | **< 0.0001** | **4.37 to 13.23** |
| Pneumothorax | **5(3.14)** | **1(0.26)** | **6(1.09)** | **0.0033** | **0.73 to 6.88** |
| CV Stroke | **2(1.26)** | **0** | **2(0.36)** | **0.22** | **-0.64 to 4.12** |
| **Outcomes** |  |  |  |  |  |
| Intubated (at least once) | 34(21.39) | 0 | 34(6.19) | < 0.0001 | 15.64 to 28.39 |
| Extubated (at least once) | 5(3.14) | 0 | 5(0.91) | 0.0004 | 1.10 to 7.14 |
| Median Average Length of Hospital Stay in Days | 11(0 to 69) | 6 (0 to 16) | 8.5 (0 to 69) | NA | NA |
| **DEATH** | 29(18.24) | 1(0.25) | 30(5.46) | 0.0001 | 12.63 to 24.72 |
| Male | 23(14.47) | 1(0.25) | 24 | <0.0001 | 9.43 to 20.52 |
| Female | 6(3.77) | 0 | 6 | 0.0001 | 1.51 to 7.98 |
| **ALIVE** | 116(72.96) | 386 (98.97) | 502(91.43) | < 0.0001 | 19.51 to 33.42 |
| **LEAVE AGAINST**<br>**MEDICAL**<br>**ADVISE(LAMA)** | 14(8.81) | 3(0.76) | 17(3.09) | < 0.0001 | 19.51 to 33.42 |

### Hospital Outcomes

Out of 549 patients, 502 (91.43%) were discharged alive; 3.1% took LAMA placing death at 5.47 % (Table 5). Of the ICU admitted patients 18.24 % succumbed to COVID with higher mortality among males in comparison to females (p<0.0001). Multivariate logistic regression analysis revealed that hypertension (OR, 1.961; 95% confidence interval, 0.80 to 4.794; p=0.14), preexisting chronic kidney diseases (OR: 6.10; p=0.02) remained to be the predictors for high mortality of COVID-19. Of the 7 ICU patients who received convalescent plasma therapy, 4 died.

## DISCUSSION

Although India is severely affected by SARS CoV infection, real world data of Indian COVID patients is inadequately available. This is the first manuscript with a data of 549 real world COVID patients at a single private center. The center hospitalized 50:50 ratio of COVID positive patients privately getting admitted as well transferred from Ahmedabad Municipal Corporation designated government hospitals. The policies, protocols, procedures were dynamic and with global literature, updates and advisories, treatment and management protocols were amended to improve patient care and outcomes.^[5,6]^ During the study period, Ministry of health and family welfare (MoHFW), India declared lockdown and encouraged mildly symptomatic patients to stay home, and isolate themselves and implemented triaging practices (including cough/cold/fever clinics, initial evaluation in primary health centers, Dhanvantri clinics to manage patients without severe dyspnea at home.^[2]^ Thus, the patients who tested positive at CIMS likely represented a higher acuity subset of symptomatic cases lately. Through manually abstracted data, this retrospective study provides an in-depth description of Indian patients with COVID-19 at a more granular level than prior literature. We hope a better understanding of our patient population, baseline characteristics, inpatient course, and clinical outcomes of Indian patients can provide valuable guidance to clinicians who are working in a time of unparalleled volume and uncertainty.

With a total of 1291 COVID positive patient admissions from 5^th^ May to 10^th^ August 2020, data of randomly 549 COVID 19 patients who received inpatient care at CIMS hospital, we found age a contributing factor to disease causation adversely affecting disease progression and outcomes. With increase in age, poorer outcomes were observed. Previous studies have mentioned that older COVID-19 patients are at an increased risk of death. ^[7-10]^

The results of a meta-analysis indicated that male sex seems to be a risk factor for mortality (both in the general population and hospitalized patients); for a lower recovery rate and for disease severity in COVID-19.^[11]^ Our data analysis highlighted a striking difference between male and female gender regarding disease susceptibility: a male to female ratio of 1:0.39, inclining towards male was observed. This striking difference depicting higher proneness of men to COVID-19 could be related to differences in innate immunity, steroid hormones and factors related to sex chromosomes crucial in the defense against viral infections. ^[12]^

Like many studies, our patients had greater prevalence of hypertension and diabetes while few had no major comorbidity. All ICU patients had one or more than one comorbidity. The findings reported here are similar with the current knowledge that the elderly and those with comorbidities are more susceptible to severe infection (Centers for Disease Control and Prevention ^[13]^ WHO-China Joint Mission. ^[14]^ A previous systematic review of 19 prevalence studies, including 2874 patients, also found hypertension as the most common comorbidity in COVID-19 patients. ^[15]^ Our study also found similar results with high rates of these comorbidities in ICU groups. For diabetes, a meta-analysis of six studies (1527 patients) found the prevalence to be twice as high in the ICU/severe group compared to non-severe COVID-19 patients.^[16]^ On disaggregating this outcome into two (non ICU and ICU admitted), we found the prevalence of diabetes to be 1.6 times higher while hypertension 1.5 times higher in the ICU group. Consistent with Sun et al., meta-analysis of symptoms in 50,466 COVID-19 patients ^[17]^ and the WHO-China joint mission on COVID-19,^[14]^ cough, fever and fatigue were the most common symptoms found in our analysis. Dyspnea was the only symptom significantly associated with disease severity and ICU admission, alongside various comorbidities (COPD, diabetes, CVD and hypertension). Patients with dyspnea were 3.4 times more likely to have an ICU admission compared to those without dyspnea. Although COPD was relatively uncommon, even in ICU patients, it was by far the most strongly predictive comorbidity for ICU admission. The significant association of dyspnea in this analysis with ICU admission suggests that silent hypoxia was less common. It should be noted though, the reported strength of dyspnea as a predictor for ICU admission.

Radiological evaluation is one way of looking in the body. ^[18]^ Chest X rays are the most commonly performed investigation in COVID 19 suspected cases.^[19]^ Presence of opacities or consolidations in the chest X ray was considered as abnormal. Although all hospitalized patients depicted positive RT-PCR, abnormal chest X rays and CT scans were observed in 56 % and 34 % of overall subjects. ICU patients were 1.9 times more likely to have an abnormal chest X –ray. Although CT scan is the preferred imaging modality regarding early detection of disease as well as of its complications but it has infection control challenges including strict decontamination measures, ventilation and airflow. ^[20]^ At our hospital, CT scan investigations were incorporated in the diagnostic protocol. The CT scan abnormalities in ICU patients were 1.5 times more than non ICU patients.

Determination of circulating D-dimer concentrations in COVID-19 patients might help to rapidly identify patients with high disease severity. Our observation of elevated D-dimer levels in ICU patients justifies it and is similar to a systematic review and meta-analysis showing that serum D-dimer concentrations in patients with severe COVID-19 are significantly higher when compared to those with non-severe forms. ^[21]^ Ferritin is a key mediator of immune dysregulation and serum ferritin levels were closely related to the severity of COVID-19. ^[22]^ In the present study overall 50% of females exhibited higher ferritin levels.

A network meta-analysis of fifteen randomized controlled trials including 8654 participants reported use of glucocorticoids, hydroxychloroquine, lopinavir-ritonavir, remdesivir, umifenovir, and standard care. ^[23]^Treatment protocols were updated with latest global information. However, hydoxychloroquine, azithromycin and steroids were prescribed in more than 50% of all patients more so in patients admitted in May and June 2020. Steroids were prescribed 2.5 times more in ICU patients than non ICU patients in consideration of increased inflammatory markers. Nearly 90 % patients depicted elevated levels of CRP. Earlier methylprednisolone was used, later replaced with dexamethasone with amendments in ICMR guidelines.^[6]^ Steroids were recommended for both preventive and treatment for cytokine storm. Remdesivir is the only intervention in which moderate certainty exists supporting benefits for both time to symptom resolution and duration of mechanical ventilation, but it remains uncertain whether remdesivir has any effect on mortality and other outcomes important to patients.^[23]^ In our study subjects, with availability of remdesivir in India, it was given to 84 % of patients more so ICU patients. Usually it was initiated between day 3 –day 5 of admission. In the present study tocilizumab (to 4-8mg/kg body weight) was administered to ICU patients. With no guidelines available the regime was used to treat patients with cytokine storm as depicted by elevated interleukin levels (data not shown). As per the first meta-analysis investigating the effect of tocilizumab on clinical outcomes of patients with severe COVID-19 mortality rate of patients treated with tocilizumab was lower than that of the control group, the difference did not reach statistical significance.^[24]^ This observation needs to be validated in comparison to the present study. Convalescent plasma therapy was also given to 7 ICU patients after ethical approval. However conclusive observations of its efficacy were not clear since 4 of these patients died.

Emerging data suggest that COVID-19 affects multiple organs, leading to organ failure and eventually death.^[25]^ Cardiovascular complications, adverse renal manifestations such as acute kidney injury (AKI), are associated with severe COVID-19 or mortality.^[26]^ As compared with prior literature, we found lower renal complications in our study subjects. Previous studies from China reported 15% of all COVID-19 patients developed AKI,^[27]^ while a case series in Seattle found 19.1% developed AKI.^[25]^. However, we found 4.37% all COVID-19 patients and 13.84% of ICU patients developed AKI, a striking decrease compared to previous reports. Concomitantly, 1.2% of all patients and 3.77% of ICU patients required inpatient hemodialysis. Shock and sepsis/secondary bacterial infection was another complication of ICU patients. At this point, the cause of the sepsis or secondary bacterial infection is difficult to ascertain since its correlation to use of tocilizumab or intubation needs to be determined.

This cohort had a median LOS of 8.5 days, which increased to 11 days for ICU patients. The overall LOS was comparable to two cohorts in China with median LOS of 11 and 12 days respectively ^[8, 28]^ One patient in ICU had a 69 days hospital stay which significantly widened the median range. With updated treatment plans the median LOS will continue to reduce given that protocols are established.

Across all levels of care,5.46 % of patients died, which is much lower to NYU (18.5%) and lies in between estimates from China (1.4%, 28%).^[29]^ Our current mortality rate of ICU patients is 18.24 %, while prior reports have suggested highly variable mortality rates in Italy (26%), China (38%, 78%), and Seattle (50%,67%).^[25]^

The results have made us realize the impact of a multi-specialty COVID care team which has a profound impact on the prognosis which is significantly dependent on baseline characteristics and investigational reports. It also envisages updating of treatment protocols based on evidenced literature within regulatory framework. The study foresees planning of post –ICU care including home needs and resuming baseline functional status.

### Study Limitations

The first limitation of the study is that data collection is limited to what is documented in the medical record data which unintentionally may be with errors in both patient recall and clinician documentation limiting accuracy of data abstraction. This potential error was minimalized by implementing a series of checks in the data. Data is representation of a single, urban medical center and may not be generalizable to all other regions. A deliberate focus was on characterizing the data of Indian patients to provide descriptive statistics and figures rather than hypothesis driven statistical inference.

## Conclusion

- A multi-specialty COVID care team improves patient outcomes.
- Baseline characteristics predict disease outcomes.
- Higher disease incidence in patients over 65 years with heavier clinical manifestations, greater severity and longer disease courses compared with those below 65 years. Closer monitoring and more medical interventions may be needed for the elder.
- Patients with high baseline comorbidities like hypertension and diabetes, developed more complications with longer hospital stays and poor outcomes.
- Males are more susceptible to SARS-CoV2 infection (M:F ratio 2.6: 1) and hold a higher chance to present with a more severe disease and poorer outcomes (4:1).
- The ICU group was older, with significantly higher proportion of males compared to the non-ICU group; and dyspnea as a major clinical characteristic and consolidation in lungs as a radiological finding.
- Treatment protocols were dynamic. Steroids, tocilizumab, remdesivir and convalescent plasma therapy were administered to ICU patients.
- Complications of acute kidney injury, shock and sepsis prevailed in ICU patients which were much lower than global rates.

## Data Availability

The data will be available from medical record software.

http://portal.cims.co/portal/login/

## Notes

### Competing Interest Statement

The authors have declared no competing interest.

### Clinical Trial

CTRI/2020/05/025247

### Funding Statement

No specific funding/grant from any agency in the public, commercial, or not-for-profit, sectors was received for this study. No funding organization or sponsor was involved in the design and conduct of the study; collection, management, analysis, and interpretation of the data; preparation, review, or approval of the manuscript; and decision to submit the manuscript for publication.

### Author Declarations

Protocol of this study was approved by the Ethics committee of CIMS hospital an Institutional Review Board and registered with Clinical trial registry of India (CTRI) no.: CTRI/2020/05/025247.

